# Latino children’s obesity risk varies by place of birth: Findings from New York City public school youth, 2006-2017

**DOI:** 10.1101/2021.03.24.21254257

**Authors:** Karen R. Flórez, Sophia E. Day, Terry T.-K. Huang, Kevin J. Konty, Emily M. D’Agostino

**Affiliations:** Center for Systems and Community Design, Graduate School of Public Health and Heath Policy, City University of New York, New York, NY; NYC Department of Health and Mental Hygiene, Office of School Health, Queens, NY; Department of Family Medicine & Community Health, Duke University School of Medicine, Durham, NC

## Abstract

**Introduction:** Research showing Place of Birth (POB) predicts excess weight gain and obesity risk among Latino adults has not prompted similar research in Latino children, although childhood is a critical period for preventing obesity.

**Methods:** Longitudinal cohort observational study on public school children self-identified by parent/guardian as Latino in grades K-12 for school years 2006-07 through 2016-17 with measured weight and height (n= 570,172_students_; 3,10,3642_observations_). POB reported by parent/guardian was categorized as continental US (not NYC) (n=295,693), NYC (n=166,361), South America (n=19,452), Central America (n=10,241), Dominican Republic (n=57,0880), Puerto Rico (n=9,687) and Mexico (n=9,647). Age- and sex-specific BMI percentiles were estimated based on established growth charts. Data were analyzed in 2020.

**Results:** Prevalence of obesity was highest among US (non-NYC)-born girls (21%) and boys (27%), followed by NYC-born girls (19%) and boys (25%). Among girls, South Americans (9%) had the lowest prevalence of all levels of obesity, while Puerto Ricans (19%) and Dominicans (15%) had the highest prevalence. Among boys, South Americans also had the lowest prevalence of all levels of obesity (15%), while Puerto Ricans (22%) and Mexicans (21%) had the highest. In adjusted models, obesity risk was highest in US (non-NYC)-born children, followed by children born in NYC (p<0.001). Immigrant Latino children exhibited an advantage, particularly Dominicans, South Americans and Puerto Ricans, and even after controlling for individual and neighborhood sociodemographic features including linguistic isolation and poverty.

**Conclusions:** The heterogeneity of obesity risk among Latino children highlights the importance of POB.

## Introduction

Childhood obesity in the U.S. has been increasing since the 1980s, and Latino children continue to be disproportionately affected.^1^ The latest national estimates from 2013-2015 show that Latino youth aged 2 to 19 years have a 23.6% prevalence of obesity and 8.4% prevalence of extreme obesity, compared to 14.7% and 4.4%, respectively, among their non-Latino white counterparts.^2-4^ Widening disparities nationwide have been documented among Latino children relative to white children.^5^ At the same time Latino children account for 25.6% of the US child population, and childhood obesity is predictive of a myriad of chronic health conditions in adulthood including asthma, arthritis, and poorer cardiometabolic and psychological risk profiles.^6-8^

Research in the U.S. has typically characterized obesity prevalence among Latino children by comparing them in aggregate to other racial/ethnic groups.^1, 2, 5^ This is despite research documenting a Latino “immigrant advantage,” or better-than-expected obesity profiles of immigrants versus those born in the U.S., despite lower SES profiles.^9-11^ Obesity research among adults has also highlighted the importance of Place of Birth (POB),^12^ including genetic, environmental, and socio-cultural factors^13, 14^ that may further drive subgroup differences among Latino children. Mexican American children have demonstrated lower obesity prevalence among children born in Mexico versus the U.S., particularly among girls.^15-17^ Other research on Latino children corroborates sex differences in obesity risk irrespective of POB.^18, 19^ To date, limited work has studied obesity prevalence among large and diverse samples of Latino children by POB and sex.

In New York City (NYC), one of the most urban and diverse areas in the U.S., statistically significant decreases are reported in the prevalence of obesity among Latino children, from 21.9% in 2006-07 to 20.2% in 2016-17.^20^ Yet, significant disparities remain among Latinos relative to their non-Latino white counterparts despite a comprehensive approach to curb obesity by the NYC Departments of Health and Mental Hygiene (DOHMH) and Education (DOE) among vulnerable communities of color.^21-23^ In this paper, we examined heterogeneity in obesity risk among Latino children attending NYC public schools by POB, while accounting for important individual and neighborhood level SES and sociocultural factors associated with obesity risk among Latino adults.^24, 25^ Identifying differences in obesity risk among Latino children by POB is important for forecasting trends in prevalence and related outcomes among this growing population.

## Methods

### Participants

Data for this study were drawn from the NYC Fitnessgram dataset jointly managed by NYC Department of Education (DOE) and DOHMH, and have been described elsewhere.^20, 26-29^ Briefly, teachers collected child-level student height and weight annually in NYC public schools. Students’ height and weight measurements were taken annually during physical education classes among K-12 students as part of the NYC Fitnessgram curriculum using a standard protocol. The study population included all NYC Latino students enrolled in a general education public school during the 2006-07 through 2016-17 school years with at least one year of height and weight measurements. Age was defined as student age on December 31^st^ of the given school year based on NYC Fitnessgram measurements that are collected annually within 3 months of this date based on testing schedules.^30^

All child-level student demographic data were drawn from NYC DOE student enrollment records linked to Fitnessgram data by a unique identifier. The DOE uses demographic information, including Latino ethnicity, only for programmatic, instructional, and administrative planning and decision-making.^31^ Parents and guardians were asked to identify whether the child was “Hispanic” (defined as “a person of Mexican, Puerto Rican, Cuban, Central or South American, or other Spanish origin”) regardless of race. Hereafter, we use the term Latino since our sample included children from, or descended from, Spanish-speaking people from Latin America. This study was approved by the City University of New York (IRB File #2015–0582) and DOHMH (Protocol # 14–019) Institutional Review Boards.

## Measures

### Independent Variable

Parents/guardians reported all demographics, including child POB, which was used to derive seven categories: continental U.S. (not NYC or U.S. territories, including Puerto Rico) (n=295,693 unique students), NYC (n=166,361 unique students), South America (n=19,452 unique students), Central America (n=10,241 unique students), Dominican Republic (n=57,0880 unique students), and Mexico (n=9,647 unique students). These subgroups were selected based on prior literature documenting important POB differences in obesity risk in national samples of adults and NYC.^12, 33^ These documented differences also drove the decision to separate Puerto Rican children from their continental U.S. counterparts (n=9,687), despite the fact that Puerto Rican children are all U.S.-born by definition. Cuban children (n=282) were excluded due to insufficient sample size to generate reliable estimates.

### Dependent Variable

The primary outcome of interest was age- and sex-specific BMI percentiles in accordance with the Centers for Disease Control and Prevention (CDC) ^34^ growth charts.^35^ Baseline age in months was calculated from the measurement date and students’ date of birth was drawn from school enrollment records. Extreme or biologically implausible values (BIV) were identified for height, weight, weight-for-height, and BMI using CDC’s age- and sex-specific criteria.^34^ An observation identified as BIV for a student in a single school year was excluded only for that school year (n=1,756 observations). The final analytic sample included 570,172 students or 3,10,3642 observations from 2006-07 through 2016-17 school years, with individual children having 1 to 11 repeated annual observations and 56% of the sample having at least 5 repeated annual observations. Weight status was defined according to CDC’s growth chart-derived norms for sex and age in months and used to compute the BMI percentile for each child as follows; underweight (BMI<5^th^ percentile), normal (5^th^ percentile≤ BMI<85th percentile), overweight (85^th^ percentile≤ BMI< 95th), obese (BMI≥ 95^th^ percentile). Obesity was stratified in accordance with previous reports^34, 36^ as follows; class I (BMI ≥95th percentile), class II (BMI >120% of the 95th percentile for age and sex or a BMI of ≥35), class III (BMI ≥140% of the 95th percentile for age and sex or a BMI of ≥40 or greater).

### Covariates

To categorize students in terms of SES, individual student household poverty (high vs. low) was based on student eligibility/non-eligibility for free/reduced-price school meals through the National School Lunch Program, which provides meal assistance according to household income at or below 185% of the federal poverty level.^37^ Area-based SES was defined according to the American Community Survey (ACS) 2012-2016 data as the percentage of households in the student’s home zip code receiving food aid from the Supplemental Nutrition Assistance Program.^38^ Neighborhood linguistic isolation was included given its significant association with BMI and obesity-related outcomes among Latinos in NYC;^25, 39^ it was measured based on the percentage of Spanish-speaking only households in the home zip code drawing from the ACS 2012–2016 and then categorized into quartiles.

### Statistical Analyses

Data were analyzed in 2020. Descriptive statistics were computed to summarize sample characteristics. Means and SDs were generated for all continuous data (BMI percentile, area poverty, and linguistic isolation), while categorical data (sex, POB, weight category, household poverty status, categorized area poverty and linguistic isolation) were reported as frequencies and percentages for all students. Continuous variables were also generated across POB, and weight categories were computed by sex and POB. Chi-square tests of homogeneity were performed to compare the distribution of weight categories across sex and POB subgroups.

Next, repeated measures mixed models were used, where repeated observations were nested within individual children who in turn were nested within home zip codes. These models estimated the association between individual-level POB and BMI percentile over time, taking into account individual- and area-level factors for all students, and also stratified by sex. Additional models were run to test whether slopes in BMI percentile over time (calendar year) were significantly different across the POB subgroups. For all models, random intercepts for area effects (home zip code) and random slopes for time point of observation (school year, to examine period cohort effects) were included with participants and zip codes as the subjects for levels 2 and 3, respectively, to account for between-child and between-zip code variations. A first-order autoregressive covariance structure was used to represent the correlated repeated measurements over time within participants in all models.

Adjusted models included sex (unstratified models), baseline age at time of BMI measurement (continuous variable), household poverty status (binary variable), home area poverty (categorical variable), linguistic isolation (categorical variable), and time (an integer value increasing from 0 to 10 corresponding to the number of repeated observations or years that each child was observed in the dataset) as covariates. Models testing whether slopes in BMI percentile over time (calendar year) were significantly different across the POB subgroups also included calendar year as a categorical variable in the models. A *p-*value of <0.05 was considered statistically significant, and all *p-*values were 2-sided. Statistical analyses were performed using SAS v.9.4.^40^

## Results

Descriptive characteristics for Latino children in grades K-12 (n_children_=809,418; n_observations_=3,103,642) appear in Table 1. The majority, 52%, of the analytic sample was born in the continental US or NYC (29%). Of the children born outside the continental US, most were from Dominican Republic (10%), followed by South America (3%), Puerto Rico (2%), Mexico (2%), and Central America (2%). There was an equal proportion of females and males, with a high household (81%) and neighborhood (44%) poverty rate, and half of the sample resided in areas with high (quartile 3) or very high (quartile 4) linguistic isolation. More than half of the children (57%) were in the healthy weight category, 21% experienced overweight, 19% were in one of the three obesity categories, and 2% experienced underweight.

**Table 1.**
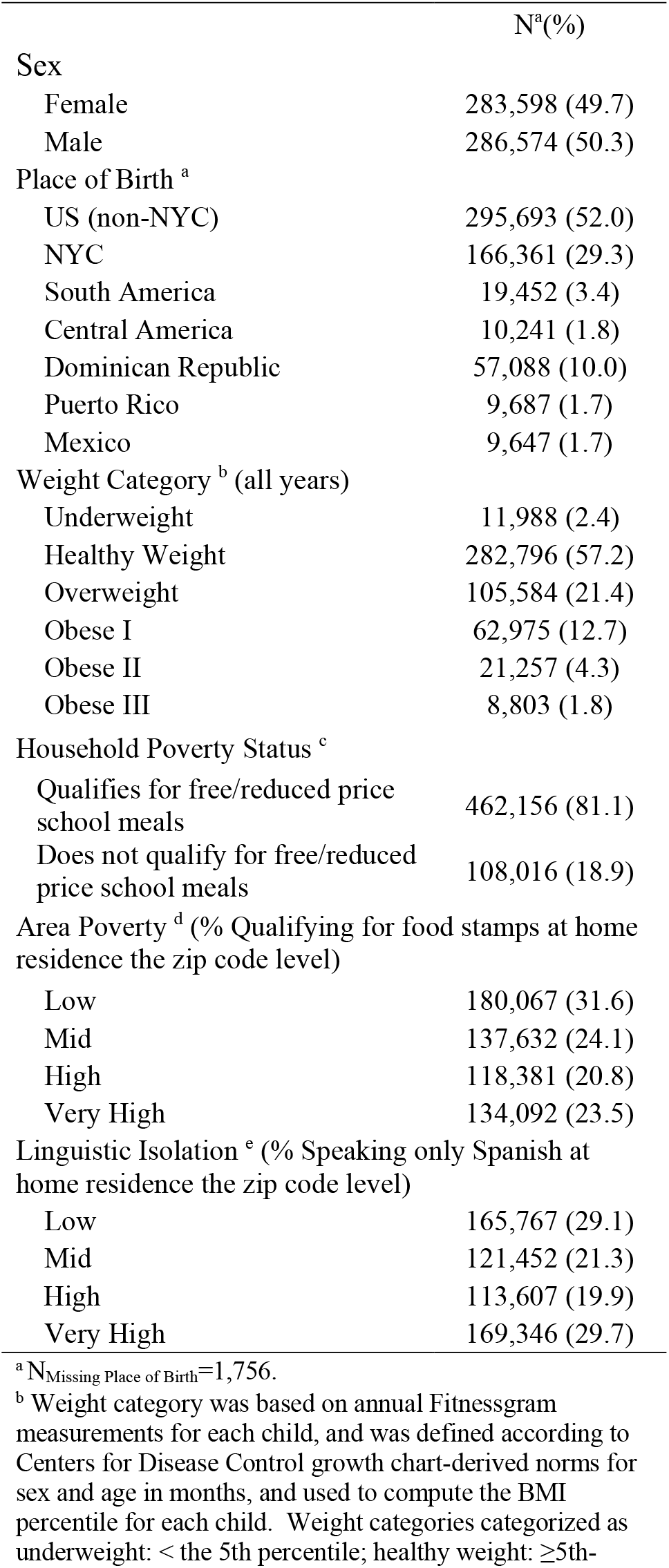
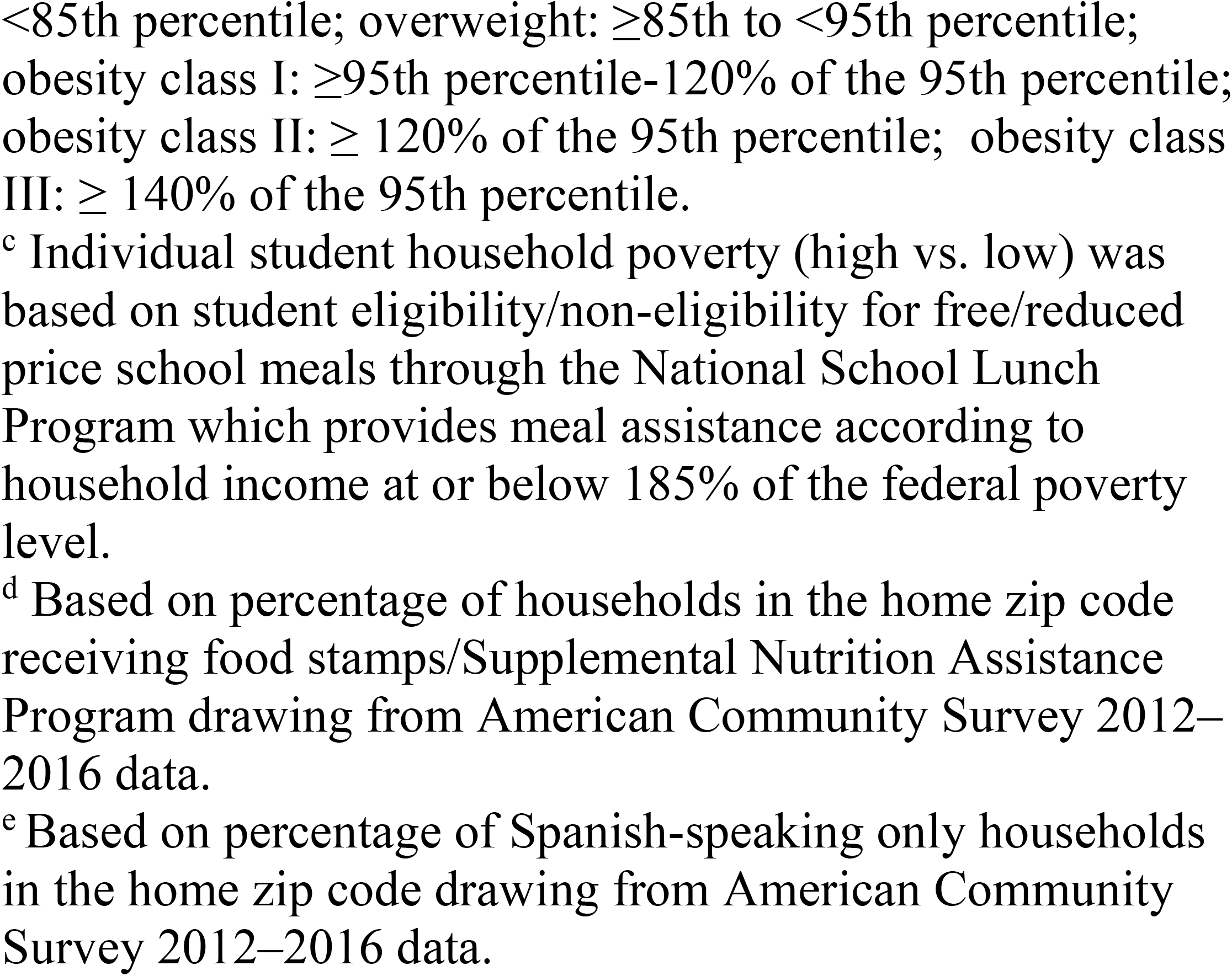
Descriptive characteristics for Latino NYC public school students (n_students_=570,172; n_observations_=3,103,642), 2006/07-2016/17

|  | N<sup>a</sup>(%) |
| --- | --- |
| Sex |  |
| Female | 283,598 (49.7) |
| Male | 286,574 (50.3) |
| Place of Birth <sup>a</sup> |  |
| US (non-NYC) | 295,693 (52.0) |
| NYC | 166,361 (29.3) |
| South America | 19,452 (3.4) |
| Central America | 10,241 (1.8) |
| Dominican Republic | 57,088 (10.0) |
| Puerto Rico | 9,687 (1.7) |
| Mexico | 9,647 (1.7) |
| Weight Category <sup>b</sup> (all years) |  |
| Underweight | 11,988 (2.4) |
| Healthy Weight | 282,796 (57.2) |
| Overweight | 105,584 (21.4) |
| Obese I | 62,975 (12.7) |
| Obese II | 21,257 (4.3) |
| Obese III | 8,803 (1.8) |
| Household Poverty Status <sup>c</sup> |  |
| Qualifies for free/reduced price school meals | 462,156 (81.1) |
| Does not qualify for free/reduced price school meals | 108,016 (18.9) |
| Area Poverty <sup>d</sup> (% Qualifying for food stamps at home residence the zip code level) |  |
| Low | 180,067 (31.6) |
| Mid | 137,632 (24.1) |
| High | 118,381 (20.8) |
| Very High | 134,092 (23.5) |
| Linguistic Isolation <sup>e</sup> (% Speaking only Spanish at home residence the zip code level) |  |
| Low | 165,767 (29.1) |
| Mid | 121,452 (21.3) |
| High | 113,607 (19.9) |
| Very High | 169,346 (29.7) |

<sup>a</sup> N<sub>Missing Place of Birth</sub>=1,756.
<sup>b</sup> Weight category was based on annual Fitnessgram measurements for each child, and was defined according to Centers for Disease Control growth chart-derived norms for sex and age in months, and used to compute the BMI percentile for each child. Weight categories categorized as underweight: < the 5th percentile; healthy weight: ≥5th-
<85th percentile; overweight: ≥ 85th to <95th percentile; obesity class I: ≥ 95th percentile-120% of the 95th percentile; obesity class II: ≥ 120% of the 95th percentile; obesity class III: ≥ 140% of the 95th percentile.
<sup>c</sup> Individual student household poverty (high vs. low) was based on student eligibility/non-eligibility for free/reduced price school meals through the National School Lunch Program which provides meal assistance according to household income at or below 185% of the federal poverty level.
<sup>d</sup> Based on percentage of households in the home zip code receiving food stamps/Supplemental Nutrition Assistance Program drawing from American Community Survey 2012–2016 data.
<sup>e</sup> Based on percentage of Spanish-speaking only households in the home zip code drawing from American Community Survey 2012–2016 data.

Supplemental Figure 1-2 and Table 2 illustrate the six weight categories for all children by POB and sex, taking into account Fitnessgram measurements collected annually from each child. Among Latina girls born in the continental US but not NYC, 21% experienced overweight, 14% were affected by Class I obesity, 5% were affected by Class II, and 2% were affected by Class III. NYC-born girls showed similar prevalence rates for all obesity categories (13%, 4%, and 2%, respectively). Girls born outside the US had a similar or lower proportion of overweight (not including youth with obesity) compared with those in the US, except for Mexican (24%) and Central American girls (22%). South and Central American, Dominican, Puerto Rican, and Mexican girls had a lower proportion of obesity in all categories compared to their US and NYC-born counterparts except for Classes I and II obesity in Puerto Rican girls (13% and 4%, respectively), which was consistent with NYC-born girls.

**Table 2.** Weight category for NYC Latino public school students, by place of birth and sex, 2006/07-2016/17.

| **Place of Birth** | **US (non-NYC)** | **NYC** | **South America** | **Central America** | **Dominican Republic** | **Puerto Rico** | **Mexico** |
| --- | --- | --- | --- | --- | --- | --- | --- |
| **Girls** | (n<sub>obs</sub>=815,285) | (n<sub>obs</sub>=494,589) | (n<sub>obs</sub>=42,910) | (n<sub>obs</sub>=18,062) | (n<sub>obs</sub>=112,670) | (n<sub>obs</sub>=21,697) | (n<sub>obs</sub>=29,622) |
| Weight Category<sup>abc</sup> | n (%) | n (%) | n (%) | n (%) | n (%) | n (%) | n (%) |
| Underweight | 19455 (2.4) | 11988 (2.4) | 941 (2.2) | 379 (2.1) | 3396 (2.8) | 539 (2.5) | 559 (1.9) |
| Healthy Weight | 453121 (55.6) | 282796 (57.2) | 29072 (67.8) | 11208 (62.1) | 75122 (61.2) | 12895 (59.4) | 17899 (60.4) |
| Overweight | 173929 (21.3) | 105584 (21.4) | 8694 (20.3) | 3995 (22.1) | 25709 (21.0) | 4135 (19.1) | 7192 (24.3) |
| Obese I | 113927 (14.0) | 62975 (12.7) | 3214 (7.5) | 1788 (9.9) | 13143 (10.7) | 2782 (12.8) | 3077 (10.4) |
| Obese II | 38689 (4.8) | 21257 (4.3) | 647 (1.5) | 463 (2.6) | 3790 (3.1) | 917 (4.2) | 578 (2.0) |
| Obese III | 14329 (1.8) | 8803 (1.8) | 188 (0.4) | 167 (0.9) | 1230 (1.0) | 343 (1.6) | 189 (0.6) |
| **Boys** | (n<sub>obs</sub>=831,849) | (n<sub>obs</sub>=498,349) | (n<sub>obs</sub>=43,326) | (n<sub>obs</sub>=19,951) | (n<sub>obs</sub>=124,104) | (n<sub>obs</sub>=20,985) | (n<sub>obs</sub>=30,243) |
| Weight Category | n (%) | n (%) | n (%) | n (%) | n (%) | n (%) | n (%) |
| Underweight | 25903 (3.1) | 15929 (3.2) | 1157 (2.7) | 472 (2.4) | 4280 (3.5) | 764 (3.6) | 882 (2.9) |
| Healthy Weight | 422248 (50.8) | 262278 (52.6) | 26687 (61.6) | 11953 (59.9) | 71660 (57.7) | 11832 (56.4) | 16678 (55.2) |
| Overweight | 159349 (19.2) | 95736 (19.2) | 8846 (20.4) | 4003 (20.1) | 23699 (19.1) | 3739 (17.8) | 6410 (21.2) |
| Obese I | 147206 (17.7) | 82589 (16.6) | 5080 (11.7) | 2512 (12.6) | 17192 (13.9) | 2912 (13.9) | 4767 (15.8) |
| Obese II | 55358 (6.7) | 30057 (6.0) | 1137 (2.6) | 715 (3.4) | 5358 (4.3) | 1180 (5.6) | 1202 (4.0) |
| Obese III | 19862 (2.4) | 10631 (2.1) | 307 (0.7) | 218 (1.1) | 1610 (1.3) | 503 (2.4) | 262 (0.9) |

<sup>a</sup> Missing weight category, Boys: US (non-NYC) n=359,307; NYC n=290,524; South America n=23,832; Central America n=13,012; Dominican Republic n=77,498; Puerto Rico n=18,720; Mexico n=21,848; Girls: US (non-NYC) n=311,662; NYC n=259,604; South America n=19,155; Central America n=9,680; Dominican Republic n=66,510; Puerto Rico n=17,052; Mexico n=17,111.
<sup>b</sup> Over half (56%) of students had at least five repeated annual observations (range 1 to 11).
<sup>c</sup> Weight category was defined according to Centers for Disease Control growth chart-derived norms for sex and age in months, and used to compute the BMI percentile for each child. Weight categories categorized as underweight: < the 5th percentile; healthy weight: ≥5th-<85th percentile; overweight: ≥85th to <95th percentile; obesity class I: ≥95th percentile-120% of the 95th percentile; obesity class II: ≥ 120% of the 95th percentile; obesity class III: ≥ 140% of the 95th percentile.

Latino boys (Supplemental figure 2 Table 2) had a higher proportion of overweight and obesity compared to girls (chi-square test comparing weight categories across POB p<.001 for all). Specifically, among Latino boys born in the continental US but not NYC, 19% experienced overweight, and 18%, 7%, and 2% experienced Classes I - III obesity, respectively. Boys born in NYC showed comparable prevalence for all obesity categories (17%, 6%, and 2%, respectively). Among children born outside the US, boys generally had a higher proportion of overweight but a lower proportion of Class I obesity (ranging from 12-16%) and Class II (ranging from 3-4%) than girls. The sole exception was Puerto Ricans, where boys had lower overweight (17.8%) than girls (19.1%) but higher Class II obesity (5.6%) than girls (4.2%). Puerto Rican boys also had a comparable prevalence of Class III compared to continental US- or NYC-born children (∼2%). All other subgroups born outside the continental US or NYC had ∼1% of boys in the Class III category.

Crude and adjusted repeated measures mixed models for the association between POB and BMI percentile for all children and stratified by sex are presented in Tables 3 and 4, respectively (Type III Fixed Effects Estimates p<.001 for all). Models adjusted for sex (unstratified model), baseline age at time of BMI measurement, household poverty status, home area poverty, linguistic isolation, calendar year, and time as covariates showed the strongest association between POB and BMI percentile in Dominican (b=-3.67, 95%CI: −3.79, −3.56), South American (b=-3.21, 95%CI: −3.40, −3.02) and Puerto Rican (b=-3.15, 95%CI: −3.44, −2.87) children, relative to the reference group (US, non-NYC-born; Table 3). After stratifying by sex, Dominican (b=-3.42, 95%CI: −3.58, −3.26), and South American (b=-3.24, 95%CI: −3.51, −2.98) girls demonstrated significantly lower BMI percentile than US non-NYC-born girls. Among boys, Puerto Ricans (b=-4.18, 95%CI: −4.59, −3.78), Dominicans (b=-3.94, 95%CI: −4.11, −3.78), Central Americans (b=-3.39, 95%CI: −3.73, −3.04) and South Americans (b=-3.19, 95%CI: −3.46, −2.92) all showed lower BMI percentile than US non-NYC born children. Also, the largest disparities by sex within the same POB subgroup were observed in Central American (b=-1.94, 95%CI: −2.30, −1.58 vs. b=-3.39, 95%CI: −3.73, −3.04) and Puerto Rican (b=-2.16, 95%CI: −2.56, −1.78 vs. b=-4.18, 95%CI: −4.59, −3.78) girls and boys, respectively.

**Table 3.** Crude and adjusted models for the association between place of birth and BMI percentile, 2006/07-2016/17.

| Place of Birth | Crude<br>Model 1 |  |  | Adjusted<br>Model 2 |  |  |
| --- | --- | --- | --- | --- | --- | --- |
|  | 95% CI |  |  | 95% CI |  |  |
| | $\beta^a$ | lower | upper | $\beta^{ab}$ | lower | upper |
| NYC | **-1.10** | -1.17 | -1.02 | **-0.67** | -0.75 | -0.59 |
| South America | **-4.10** | -4.29 | -3.92 | **-3.21** | -3.40 | -3.02 |
| Central America | **-3.31** | -3.56 | -3.06 | **-2.73** | -2.98 | -2.48 |
| Dominican Republic | **-3.79** | -3.90 | -3.67 | **-3.67** | -3.79 | -3.56 |
| Puerto Rico | **-3.32** | -3.60 | -3.04 | **-3.15** | -3.44 | -2.87 |
| Mexico | **-2.12** | -2.40 | -1.86 | **-1.23** | -1.49 | -0.97 |
| US, non-NYC | . | . | . | . | . | . |

<sup>a</sup> Abbreviations: CI, Confidence Interval; NYC, New York City; US, non-NYC, United States, not New York City.
<sup>b</sup> Boldface indicates statistical significance ( $p < 0.05$ ).
<sup>c</sup> Beta estimates generated on three-level repeated measures mixed models using SAS PROC GLIMMIX.
<sup>d</sup> Adjusted models included sex, baseline age at time of BMI measurement (continuous variable), household poverty status (binary variable), home area poverty (categorical variable), linguistic isolation (categorical variable), and time (an integer value increasing from 0 to 10 corresponding to the number of repeated observations or years that each child was observed in the dataset) as covariates.

**Table 4.** Models for the association between place of birth and BMI percentile across sex, 2006/07-2016/17.

| Place of Birth | Girls |  |  |  |  |  | Boys |  |  |  |  |  |
| --- | --- | --- | --- | --- | --- | --- | --- | --- | --- | --- | --- | --- |
|  | Crude<br>Model 3 |  |  | Adjusted<br>Model 4 |  |  | Crude<br>Model 5 |  |  | Adjusted<br>Model 6 |  |  |
|  | 95% CI |  |  | 95% CI |  |  | 95% CI |  |  | 95% CI |  |  |
| | $\beta^a$ | lower | upper | $\beta^{ab}$ | lower | upper | $\beta^a$ | lower | upper | $\beta^{ab}$ | lower | upper |
| NYC | **-0.91** | -1.02 | -0.81 | **-0.71** | -0.82 | -0.60 | **-1.30** | -1.39 | -1.17 | **-0.69** | -0.80 | -0.58 |
| South America | **-3.90** | -4.16 | -3.65 | **-3.24** | -3.51 | -2.98 | **-4.30** | -4.57 | -4.57 | **-3.19** | -3.46 | -2.92 |
| Central America | **-2.29** | -2.66 | -1.93 | **-1.94** | -2.30 | -1.58 | **-4.27** | -4.62 | -4.62 | **-3.39** | -3.73 | -3.04 |
| Dominican Republic | **-3.17** | -3.32 | -3.01 | **-3.42** | -3.58 | -3.26 | **-4.65** | -4.56 | -4.56 | **-3.94** | -4.11 | -3.78 |
| Puerto Rico | **-2.00** | -2.39 | -1.61 | **-2.16** | -2.56 | -1.78 | **-4.64** | -5.05 | -5.05 | **-4.18** | -4.59 | -3.78 |
| Mexico | **-1.82** | -2.19 | -1.46 | **-1.39** | -1.75 | -1.02 | **-2.41** | -2.78 | -2.78 | **-1.08** | -1.46 | -0.70 |
| US, non-NYC | . | . | . | . | . | . | . | . | . | . | . | . |

<sup>a</sup> Abbreviations: CI, Confidence Interval; NYC, New York City; US, non-NYC, United States, not New York City.
<sup>b</sup> Boldface indicates statistical significance ( $p < 0.05$ ).
<sup>c</sup> Beta estimates generated on three-level repeated measures mixed models using SAS PROC GLIMMIX.
<sup>d</sup> Adjusted models included baseline age at time of BMI measurement (continuous variable), household poverty status (binary variable), home area poverty (categorical variable), linguistic isolation (categorical variable), and time (an integer value increasing from 0 to 10 corresponding to the number of repeated observations or years that each child was observed in the dataset) as covariates.

Additional models examining whether slopes over time (calendar year) were significantly different across the POB subgroups showed that Puerto Rican children had the greatest decline in BMI percentile over time (b=-2.38, 95%CI: −3.31, −1.44), followed by Dominican (b=-1.99, 95%CI: −2.33, −1.65) and Central American b=-1.70, 95%CI: 2.42, −0.98) children; (Type III Fixed Effects Estimates p<.001 for all; Table S1 and Supplemental Figure 3). Models stratified by sex showed similar findings among boys (b=-2.79, 95%CI: −4.13, −1.44, b=-2.40, 95%CI: - 2.89, −1.91 and b=-2.31, 95%CI: −3.30, −1.32 for Puerto Rican, Dominican and Central American children, respectively). However, among girls, Puerto Rican (b=2.53, 95%CI: −3.83, 1.24), Dominican (b=-2.17, 95%CI: −2.65, −1.70) and South American (b=-2.07, 95%CI: −2.95, −1.19) children showed the greatest declines in BMI percentile.

## Discussion

These findings show that Latino children attending NYC public schools in grades K-12 have a high burden of excess weight and obesity risk; however, this burden is not distributed evenly across POB. Specifically, children born in the continental US had the highest proportion of excess weight and obesity risk, followed by children born in NYC. Children born outside the US did exhibit an immigrant advantage, but this was most pronounced among Dominicans, South Americans, and Puerto Ricans and, to a less degree, among Central Americans and Mexicans. This early immigrant advantage among Latino children could potentially translate into protective effects documented among immigrant adults for metabolic diseases.^10, 41, 42^ However, prevalence of obesity by POB among Latino adults revealed the highest prevalence among Puerto Ricans and the lowest among South Americans.^4^ This suggests that the early immigrant advantage observed in children may not confer the same benefits into adulthood for all Latinos, and a robust line of sociological research suggests the importance of understanding the social and economic factors that create segmented patterns of experiences by POB.^44-46^ The potential for obesity risk to follow similar segmented patterns by POB should be explored in future studies using longitudinal designs.

The literature has failed to address the heterogeneity in obesity risk among Latino children of different national or cultural origin. For example, previous surveillance data from the NYC DOHMH showed that the obesity prevalence among all Latino NYC public high school students was 17% among male and 14% among female students,^47^ masking the risk and advantage among some subgroups. This might explain why a recent study using data with grades K-8 showed that the decrease in obesity in terms of both absolute and relative prevalence remained smaller among Latino children than white children.^20^ Combining Latino children into one large category obscures relevant differences for understanding risk in excess weight and obesity in this growing segment of the population. Although research and practitioners have a growing recognition of differing health patterns and profiles among the various Latino groups, to date, there are almost no data on obesity of subgroups such as Dominicans, who constitute the fifth largest Latino group in the US.^48^ An exception is the Hispanic Community Children’s Health Study/Study of Latino Youth, though prevalence of obesity was stratified by those of Mexican background vs. Non-Mexican background.^18^

Further understanding of obesity risk among Latino children will likely require an exploration of other broad social determinants of health that might explain some of the patterns observed in this dataset. This study accounted for neighborhood-level sociodemographic and cultural factors as covariates, including area poverty derived from the proportion of residents receiving food assistance, as well as the proportion of residents speaking only Spanish in the home as a proxy for linguistic isolation. Others have pointed to a wider array of neighborhood and social factors that may be critical in obesity prevention throughout the life course,^14^ and factors that may be particularly important for Latino communities such as strategies that focus more on behavior change.^49^ Contextualizing these factors within a theoretical framework will be important to move beyond documenting differences; rather, we need to move into the next phase of research to explore the risk and resilience mechanisms that are shaped by complex sociocultural and SES factors.^50, 51^

This study has some limitations. First, the results do not include private, charter, and special education schools, which constitute around 18%, 10%, and 2% of elementary and middle school children, respectively, in NYC; since these children do not participate in NYC Fitnessgram. Second, not all NYC public school children participate in Fitnessgram every year, although since 2010 schools have been incentivized to collect data from a minimum of 85% of students who do not have a testing waiver. The majority of observations in this study came from younger children in elementary and middle school (60%), indicating that high school children disproportionately missed the NYC Fitnessgram assessment. However, the size and heterogeneity of the complete sample, and the ability to stratify analyses based on key demographic factors while maintaining statistical power, are strengths. Third, US-born and NYC-born children may live in households that are still deeply connected to countries throughout Latin America and this dataset does not allow for this exploration. Fourth, this dataset does not allow for a disaggregation of Central and South Americans.

## Conclusions

The NYC public school system is the largest in the country, serving 1.1 million children and representing all countries in Latin America.^52^ Findings presented in this study highlight important disparities by POB and sex after controlling for individual- and neighborhood-level factors. This work combats misperceptions of weight in this population and calls for interventions that promote positive attitudes and behaviors toward achieving a healthy weight.^53^ Future research should also examine factors in the U.S. that influence immigration waves, neighborhood composition, intergenerational patterns, and that may be working in tandem to exacerbate obesity *risk and/or resiliency* among Latino children in different settings.

## Data Availability

This surveillance data is not available without permission from the NYC Department of Health

## Notes

### Competing Interest Statement

The authors have declared no competing interest.

### Funding Statement

The content is solely the responsibility of the authors and does not represent the official views of NIDDK/NIH. Dr. Florez received salary support from grant 1R21DK114630-01A1 from the National Institute of Diabetes and Digestive and Kidney Diseases of the National Institutes of Health (NIDDK/NIH).

### Author Declarations

This study was approved by the City University of New York (IRB File #2015 0582)and DOHMH (Protocol # 14 019) Institutional Review Boards.

## References

1. Skinner AC, Ravanbakht SN, Skelton JA, Perrin EM, Armstrong SC. Prevalence of Obesity and Severe Obesity in US Children, 1999-2016. Pediatrics. Mar 2018;141(3).

2. Ogden CL, Carroll MD, Fakhouri TH, et al. Prevalence of Obesity Among Youths by Household Income and Education Level of Head of Household - United States 2011-2014. MMWR Morb Mortal Wkly Rep. Feb 16 2018;67(6):186–189.

3. Hales CM, Carroll MD, Fryar CD, Ogden CL. Prevalence of Obesity Among Adults and Youth: United States, 2015-2016. NCHS Data Brief. Oct 2017(288):1–8.

4. Hales CM, Fryar CD, Carroll MD, Freedman DS, Ogden CL. Trends in Obesity and Severe Obesity Prevalence in US Youth and Adults by Sex and Age, 2007-2008 to 2015-2016. JAMA. Apr 24 2018;319(16):1723–1725.

5. Isong IA, Rao SR, Bind MA, Avendano M, Kawachi I, Richmond TK. Racial and Ethnic Disparities in Early Childhood Obesity. Pediatrics. Jan 2018;141(1).

6. The NS, Suchindran C, North KE, Popkin BM, Gordon-Larsen P. Association of adolescent obesity with risk of severe obesity in adulthood. JAMA. Nov 10 2010;304(18):2042–2047.

7. Reilly JJ, Kelly J. Long-term impact of overweight and obesity in childhood and adolescence on morbidity and premature mortality in adulthood: systematic review. Int J Obes (Lond). Jul 2011;35(7):891–898.

8. US Census Bureau. The Hispanic Population in the United States. Available at: https://www.census.gov/data/tables/2019/demo/hispanic-origin/2019-cps.html. Accessed June 30, 2020.

9. Abraido-Lanza AF, Chao MT, Florez KR. Do healthy behaviors decline with greater acculturation? Implications for the Latino mortality paradox. Soc Sci Med. Sep 2005;61(6):1243–1255.

10. Abraido-Lanza AF, Echeverria SE, Florez KR. Latino Immigrants, Acculturation, and Health: Promising New Directions in Research. Annu Rev Public Health. 2016;37:219–236.

11. Chen Y, Freedman ND, Rodriquez EJ, et al. Trends in Premature Deaths Among Adults in the United States and Latin America. JAMA Netw Open. Feb 5 2020;3(2):e1921085.

12. Bates LM, Acevedo-Garcia D, Alegria M, Krieger N. Immigration and generational trends in body mass index and obesity in the United States: results of the National Latino and Asian American Survey, 2002-2003. Am J Public Health. Jan 2008;98(1):70–77.

13. Moon JY, Wang T, Sofer T, et al. Objectively Measured Physical Activity, Sedentary Behavior, and Genetic Predisposition to Obesity in U.S. Hispanics/Latinos: Results From the Hispanic Community Health Study/Study of Latinos (HCHS/SOL). Diabetes. Dec 2017;66(12):3001–3012.

14. Suglia SF, Duarte CS, Chambers EC, Boynton-Jarrett R. Cumulative social risk and obesity in early childhood. Pediatrics. May 2012;129(5):e1173–1179.

15. Winham DM. Growth status among low-income Mexican and Mexican-American elementary school children. Am J Hum Biol. Sep-Oct 2012;24(5):690–695.

16. Hernandez-Valero MA, Wilkinson AV, Forman MR, et al. Maternal BMI and country of birth as indicators of childhood obesity in children of Mexican origin. Obesity (Silver Spring). Oct 2007;15(10):2512–2519.

17. Lawrence E, Mollborn S, Riosmena F. Early Childhood Disadvantage for Sons of Mexican Immigrants: Body Mass Index Across Ages 2-5. Am J Health Promot. Sep 2016;30(7):545–553.

18. Isasi CR, Parrinello CM, Ayala GX, et al. Sex Differences in Cardiometabolic Risk Factors among Hispanic/Latino Youth. J Pediatr. Sep 2016;176:121–127 e121.

19. Isasi CR, Whiffen A, Campbell E, Florez Y, Freeman K, Wylie-Rosett J. High prevalence of obesity among inner-city adolescent boys in the Bronx, New York: forgetting our boys. Prev Chronic Dis. Jan 2011;8(1):A23.

20. Day SE, D’Agostino EM, Huang TT, Larkin M, Harr L, Konty KJ. Continued Decline in Obesity and Severe Obesity Prevalence Among New York City Public School Youth in Grades K-8: 2011-2017. Obesity (Silver Spring). Mar 2020;28(3):638–646.

21. Lederer A, Curtis CJ, Silver LD, Angell SY. Toward a healthier city: nutrition standards for New York City government. Am J Prev Med. Apr 2014;46(4):423–428.

22. Dunn LL, Venturanza JA, Walsh RJ, Nonas CA. An observational evaluation of move-to-improve, a classroom-based physical activity program, New York City schools, 2010. Prev Chronic Dis. 2012;9:E146.

23. Cawley J, Cisek-Gilman L, Roberts R, et al. Effects of HealthCorpos, a High School Mentoring Program, on Youth Diet and Physical Activity. Childhood Obesity. 2011;7(5):364–371.

24. Rundle A, Richards C, Bader MD, et al. Individual-and school-level sociodemographic predictors of obesity among New York City public school children. Am J Epidemiol. Dec 1 2012;176(11):986–994.

25. Park Y, Neckerman KM, Quinn J, Weiss C, Rundle A. Place of birth, duration of residence, neighborhood immigrant composition and body mass index in New York City. Int J Behav Nutr Phys Act. Apr 6 2008;5:19.

26. Bezold CP, Stark JH, Rundle A, et al. Relationship between Recreational Resources in the School Neighborhood and Changes in Fitness in New York City Public School Students. J Urban Health. Feb 2017;94(1):20–29.

27. D’Agostino EM, Day SE, Konty KJ, Larkin M, Saha S, Wyka K. The association of fitness and school absenteeism across gender and poverty: a prospective multilevel analysis in New York City middle schools. Ann Epidemiol. Mar 2018;28(3):189–196.

28. Day SE, Konty KJ, Leventer-Roberts M, Nonas C, Harris TG. Severe obesity among children in New York City public elementary and middle schools, school years 2006-07 through 2010-11. Prev Chronic Dis. Jul 10 2014;11:E118.

29. Centers for Disease Control Prevention. Obesity in K-8 students - New York City, 2006-07 to 2010-11 school years. MMWR Morb Mortal Wkly Rep. Dec 16 2011;60(49):1673–1678.

30. NYC Department of Education. 2019-20 NYC Fitnessgram pacing calendar grades 9-12. Available at: https://www.weteachnyc.org/resources/resource/2018-19-nyc-fitnessgram-pacing-calendar-grades-9-12-physical-best-spring-only/. Accessed January 4, 2021.

31. NYC Department of Education. Non-Discrimination Policy. Available at: https://www.schools.nyc.gov/about-us/policies/non-discrimination-policy. Accessed July 7, 2020.

32. NYC Department of Education. English Language Learners. Available at: https://www.schools.nyc.gov/learning/multilingual-learners/english-language-learners. Accessed July 7, 2020.

33. Greer S, Naidoo M, Hinterland K, et al. Health of Latinos in NYC 2017.

34. Kuczmarski RJ, Ogden CL, Grummer-Strawn LM, et al. CDC growth charts: United States. Adv Data. Jun 8 2000(314):1–27.

35. A SAS Program for the 2000 CDC growth charts (ages 0 to < 20 years) [computer program]. Version; 2019.

36. Freedman DS, Berenson GS. Tracking of BMI z Scores for Severe Obesity. Pediatrics. Sep 2017;140(3).

37. Tropini A J.L. H., Selecting and applying a standard area-based socioeconomic status measure for public health data: analysis of New York City. 2013.

38. US Census Bureau. American Community Survey (ACS). Available at: https://www.census.gov/programs-surveys/acs. Accessed June 30, 2020.

39. Park Y, Neckerman K, Quinn J, Weiss C, Jacobson J, Rundle A. Neighbourhood immigrant acculturation and diet among Hispanic female residents of New York City. Public Health Nutr. Sep 2011;14(9):1593–1600.

40. SAS version 9.4 [computer program]. Version. Cary, North Carolina.

41. Bullard KM, Cowie CC, Lessem SE, et al. Prevalence of Diagnosed Diabetes in Adults by Diabetes Type - United States, 2016. MMWR Morb Mortal Wkly Rep. Mar 30 2018;67(12):359–361.

42. Saab S, Manne V, Nieto J, Schwimmer JB, Chalasani NP. Nonalcoholic Fatty Liver Disease in Latinos. Clin Gastroenterol Hepatol. Jan 2016;14(1):5–12; quiz e19-10.

43. Isasi CR, Ayala GX, Sotres-Alvarez D, et al. Is acculturation related to obesity in Hispanic/Latino adults? Results from the Hispanic community health study/study of Latinos. J Obes. 2015;2015:186276.

44. Zhou M. Segmented assimilation: issues, controversies, and recent research on the new second generation. Int Migr Rev. Winter 1997;31(4):975-971,008.

45. Xie Y, Greenman E. The social context of assimilation: testing implications of segmented assimilation theory. Soc Sci Res. May 2011;40(3):965–984.

46. Portes A, Fernández-Kelly P, Haller W. The Adaptation of the Immigrant Second Generation in America: Theoretical Overview and Recent Evidence. J Ethn Migr Stud. 2009;35(7):1077–1104.

47. Greer S, Naidoo M, Hinterland K, et al. Health of Latinos 2017.

48. Noe-Bustamante L, Flores A, Shah S. Facts on Hispanics of Dominican origin in the United States 2017.

49. Woodward-Lopez G, Gosliner W, Au LE, et al. Community characteristics modify the relationship between obesity prevention efforts and dietary intake in children: the Healthy Communities Study. Pediatr Obes. Oct 2018;13 Suppl 1:46–55.

50. Taveras EM, Gillman MW, Kleinman KP, Rich-Edwards JW, Rifas-Shiman SL. Reducing racial/ethnic disparities in childhood obesity: the role of early life risk factors. JAMA Pediatr. Aug 1 2013;167(8):731–738.

51. Abraido-Lanza AF, Mendoza-Grey S, Florez KR. A Commentary on the Latin American Paradox. JAMA Netw Open. Feb 5 2020;3(2):e1921165.

52. NYC Department of Education. DOE Data at a Glance. Available at: https://www.schools.nyc.gov/about-us/reports/doe-data-at-a-glance. xAccessed July 7, 2020.

53. Turer CB, Montano S, Lin H, Hoang K, Flores G. Pediatricians’ communication about weight with overweight Latino children and their parents. Pediatrics. Nov 2014;134(5):892–899.

