## Supplemental Table 1 for "Latino children’s obesity risk varies by place of birth: Findings from New York City public school youth, 2006-2017"

| **Table S1**. Models for the association between place of birth*year and BMI percentile across sex, 2006/07-2016/17. | | | | | | | | | | | | | | | | | | | | | | | |
| --- | --- | --- | --- | --- | --- | --- | --- | --- | --- | --- | --- | --- | --- | --- | --- | --- | --- | --- | --- | --- | --- | --- | --- |
|  | **Girls** | | | | | | | | | | **Boys** | | | | | | | | | | | | |
| **Place of Birth** | **Crude** | | | | | **Adjusted** | | | | | **Crude** | | | | | | | | **Adjusted** | | | | |
|  | Model 1 | | | | | Model 2 | | | | | Model 1 | | | | | | | | Model 2 | | | | |
|  |  | **95% CI** | | | |  | **95% CI** | | | |  | | | **95% CI** | | | | |  | | **95% CI** | | |
|  | **β^a^** | **lower** | **upper** | | | **β^ab^** | **lower** | | **upper** | | **β^a^** | | | **lower** | | | **upper** | | **β^ab^** | | **lower** | | **upper** |
| NYC | **0.08** | -0.24 | 0.40 | | | **0.49** | 0.17 | | 0.81 | | **-0.47** | | | -0.80 | | | -0.14 | | **0.35** | | 0.03 | | 0.68 |
| South American | **-2.07** | -2.95 | -1.19 | | | **-1.26** | -2.14 | | -0.39 | | **-3.05** | | | -3.95 | | | -2.15 | | **-1.00** | | -1.89 | | -0.10 |
| Central American | **-1.73** | -2.78 | -0.69 | | | **-0.92** | -1.96 | | 0.13 | | **-4.65** | | | -5.65 | | | -3.66 | | **-2.31** | | -3.30 | | -1.32 |
| Dominican | **-2.17** | -2.65 | -1.70 | | | **-1.58** | -2.05 | | -1.10 | | **-4.02** | | | -4.51 | | | -3.53 | | **-2.40** | | -2.89 | | -1.91 |
| Puerto Rican | **-2.53** | -3.83 | -1.24 | | | **-2.05** | -3.34 | | -0.75 | | **-4.06** | | | -5.41 | | | -2.71 | | **-2.79** | | -4.13 | | -1.44 |
| Mexican | **-0.28** | -1.54 | 0.99 | | | **0.98** | -0.29 | | 2.25 | | **-3.11** | | | -4.40 | | | -1.82 | | **0.17** | | -1.12 | | 1.45 |
| US, non-NYC | . | . | . | | | . | . | | . | | . | | | . | | | . | | . | | . | | . |
| a Abbreviations: CI, Confidence Interval; NYC, New York City; US, non-NYC, United States, not New York City.  ^b^ Boldface indicates statistical significance (p<0.05). | | | | | | | | | | | | | | | | | | | | | | | |
| ^a^Beta estimates generated on three-level repeated measures mixed models using SAS PROC GLIMMIX. | | | | | | | | | | | | | | | | | | | | | | | |
| ^b^Adjusted models included age at time of BMI measurement (continuous variable), household poverty status (binary variable), home area poverty (categorical variable), linguistic isolation (categorical variable), calendar year (to account to potential cohort effects), time (an integer value increasing from 0 to 10 corresponding to the number of repeated observations or years that each child was observed in the dataset) as covariates, as well as a place of birth*calendar year interaction term. | | | | | | | | | | | | | | | | | | | | | | | |
| *All beta estimates, p<0.001. | | | | | | | | | | |  | | |  | | |  | |  | |  | |  |
