## Supplemental Figure 1 for "Latino children’s obesity risk varies by place of birth: Findings from New York City public school youth, 2006-2017"

Figure S1. Weight Category for NYC Latino public school BOYS in grades k-12 by Place of birth (nobservations=1,568,807; nchildren=286,574), 2006/07-2016/17

|  | US (non-NYC) | NYC | South America | Central America | Dominican Republic | Puerto Rico | Mexico |
| --- | --- | --- | --- | --- | --- | --- | --- |
| Underweight | 3.3 | 3.3 | 3 | 2.7 | 3.6 | 3.5 | 2.7 |
| Healthy Weight | 54.3 | 55.7 | 64.4 | 61.6 | 59.9 | 58.1 | 55.6 |
| Overweight | 19.3 | 19.7 | 20.2 | 20 | 19.6 | 17.6 | 22 |
| Obese I | 15.4 | 15.3 | 9.5 | 11.5 | 12.3 | 13.3 | 15.4 |
| Obese II | 5.4 | 5 | 2.1 | 3 | 3.4 | 5.2 | 3.5 |
| Obese III | 2.1 | 1.8 | 0.5 | 0.8 | 1 | 2.1 | 0.74 |
