## Supplemental Figure 2 for "Latino children’s obesity risk varies by place of birth: Findings from New York City public school youth, 2006-2017"

Figure S2. Weight Category for NYC Latino public school Girls in grades k-12 by Place of birth (nobservations=1,534,835; nchildren=283,598), 2006/07-2016/17

|  | US (non-NYC) | NYC | South America | Central America | Dominican Republic | Puerto Rico | Mexico |
| --- | --- | --- | --- | --- | --- | --- | --- |
| Underweight | 2.7 | 2.6 | 2.3 | 2.6 | 3.2 | 2.9 | 1.9 |
| Healthy Weight | 58.7 | 60.5 | 70.8 | 63.7 | 65.2 | 59.8 | 63.7 |
| Overweight | 21.4 | 21.3 | 19 | 21.6 | 20 | 19.8 | 23.1 |
| Obese I | 11.4 | 10.2 | 6 | 8.4 | 8.2 | 3.1 | 9.1 |
| Obese II | 3.9 | 3.5 | 1.1 | 2.2 | 2.22 | 3.1 | 1.3 |
| Obese III | 1.7 | 1.7 | 0.4 | 1.1 | 0.9 | 1.5 | 0.5 |
