## Supplemental Figure 3 for "Latino children’s obesity risk varies by place of birth: Findings from New York City public school youth, 2006-2017"

Figure S3. Association between place of birth*calendar year and body mass index percentile for NYC Latino public school students in grades K-12 (nobservations=3,103,642; nchildren=809,418) across sex, 2006/07-2016/17

|  | Change in BMI Percentile |
| --- | --- |
| Girls NYC | 0.492 |
| Girls South America | -1.262 |
| Girls Central America | -0.916 |
| Girls Dominican Republic | -1.576 |
| Girls Puerto Rican | -2.046 |
| Girls Mexican | 0.978 |
| Boys NYC | 0.354 |
| Boys South America | -0.996 |
| Boys Central America | -2.307 |
| Boys Dominican Republic | -2.398 |
| Boys Puerto Rican | -2.787 |
| Boys Mexican | 0.168 |
